# From Vaccine to Pathogen: Modeling Sabin 2 Vaccine Virus Reversion and Evolutionary Epidemiology

**DOI:** 10.1101/2020.11.02.20224634

**Authors:** Wesley Wong, Jillian Gauld, Michael Famulare

## Abstract

The oral poliovirus vaccines (OPV) are one of most effective disease eradication tools in public health. However, the Sabin 2 vaccine strain can revert attenuation and cause outbreaks of circulating, vaccine-derived poliovirus (cVDPV2) that are clinically indistinguishable from wild poliovirus (WPV). Accurately assessing cVDP2 risk requires disentangling the complex interaction between epidemiology and evolutionary biology. Here, we developed a Sabin 2 reversion model that simulates the reversion of Sabin 2 to WPV based on the clinical differences in shedding duration and infectiousness between individuals vaccinated with Sabin 2 and those infected with wild poliovirus. Genetic reversion is informed by a canonical reversion pathway defined by three gatekeeper mutations (A481G, U2909C, and U398C) and the accumulation of genetic load from deleterious nonsynonymous mutations. Our model captures essential aspects of both phenotypic and molecular evolution and simulates transmission using a multiscale transmission model that consolidates the relationships among immunity, susceptibility, and transmission risk. We show that despite the rapid reversion of Sabin 2, cVDPV2 outbreaks can be controlled by maintaining high levels of population-level immunity and sanitation. Supplementary immunization activities must maintain high vaccine coverage to prevent future cVDPV2 outbreaks in the targeted intervention zone, but declining global immunity against Sabin 2 makes them increasingly risky to implement in poor sanitation regions regardless of historical immunization activity. A combined strategy of assessing and improving sanitation levels in conjunction with high coverage vaccination campaigns will limit future cVDPV2 emergence and spread.

**Significance Statement:** Since the withdrawal of the Sabin 2 oral poliovirus vaccine (OPV2), circulating vaccine-derived poliovirus outbreaks caused by the genetic reversion of Sabin 2 vaccine virus (cVDPV2) have been increasing in frequency. The current strategies for combating cVDPV2 involve supplemental immunization activities with monovalent Sabin 2 oral poliovirus (mOPV2), which can inadvertently seed future cVDPV2 outbreaks. Accurately assessing future cVDPV2 outbreak risk following mass mOPV2 campaigns is critical poliovirus eradication efforts but must consider the interaction between genetic reversion and epidemiological transmission. We developed an evolutionary epidemiology model to integrate Sabin 2 genetic reversion and transmission into a single framework to evaluate their relative contribution to cVDPV2 outbreaks and inform future intervention strategies.

## Introduction

Mass immunization with oral poliovirus vaccine (OPV) has led to a >99.99% drop in wild poliovirus (WPV) cases (1) and the complete eradication of Type 2 and Type 3 WPV. The OPVs are highly efficacious vaccines that confer long-lasting protective immunity (2). A curious feature of these vaccines is that vaccinated individuals can shed and transmit vaccine virus. Vaccine transmission increases population-level immunity but is problematic when viruses revert attenuation, resulting in circulating, vaccine-derived poliovirus (cVDPV) capable of causing paralytic poliomyelitis cases clinically indistinguishable from WPV (3). Following the eradication of Type 2 WPV, the Global Polio Eradication Initiative coordinated the global withdrawal of Sabin 2 OPV (OPV2) from routine immunization schedules (commonly referred to as the Switch) to prevent future sources of circulating, vaccine-derived Type 2 poliovirus (cVDPV2) (1).

Despite the Switch, cVDPV2 outbreaks continue to be a pressing public health concern and have emerged as a significant issue threatening global poliovirus eradication efforts (1, 4). Although OPV2 is no longer used in routine immunization, monovalent OPV2 (mOPV2) vaccination campaigns are still used as supplemental immunization activities (SIAs) to combat cVDPV2 outbreaks. cVDPV2 outbreaks are now reported throughout the African continent, Pakistan, Afghanistan, China, Malaysia and the Philippines and 29% of the outbreaks in Africa have involved international spread (4, 5). Since the Switch, 126 mOPV2 campaigns utilizing more than 300 million doses of OPV2 have been implemented to control cVDPV2 outbreaks (6). These SIAs are estimated to be responsible for seeding over half of the cVDPV2 outbreaks observed since the Switch (6). Ensuring the future safety and efficacy of these SIA campaigns while minimizing the risk of seeding future cVDPV2 outbreaks is critical to the success of the poliovirus eradication initiative.

The Sabin 2 vaccine strain was derived from a wild-type isolate (P22/P712/56) passaged in monkey kidney cells to achieve neurovirulence attenuation (7). However, identifying the genetic mutations responsible for attenuation has been surprisingly complicated, in part because the progenitor has not been fully sequenced and because P22/P712/56 is thought to be naturally attenuated relative to other WPV strains (8). Despite this, phylogenetic analyses of whole genome sequences collected from cVDPV2 outbreaks suggest a common evolutionary pathway to attenuation reversal (9). This pathway is characterized by the rapid fixation three gatekeeper mutations, A481G, U2909C, and U398C, within the first few weeks after vaccination (9, 10). All three mutations have previously been implicated as molecular determinants of attenuation for Sabin 2 (11–14). A481G and U398C are noncoding mutations that affect the stability of a 5’ hairpin structure that mediates translation efficiency (13, 15, 16) and U2909C is a nonsynonymous mutation in the VP1 capsid protein.

However, genetic reversion is not the sole determinant of cVDPV2 emergence. Environmental sewer samples have identified revertant strains in regions with no detectable cVDPV2 cases (17). In fact, vaccinated individuals can shed and transmit revertant viruses to other individuals (10, 18), but whether this results in clinically-detected cVDPV2 outbreaks depends on the epidemiological context. These cVDPV2 outbreaks are associated with regions with historically poor vaccination rates and low population-level immunity (6, 19). Thus, while Sabin 2 reversion is a prerequisite of cVDPV2 emergence, whether it triggers a public health crisis depends on the interactions between genetic reversion and local epidemiological conditions.

Mathematical models can be useful tools for assessing cVDPV2 emergence risk and characterizing the evolutionary and epidemiological factors that allow cVDPV2 outbreaks to occur. We developed an evolutionary-epidemiology model that simulates Sabin 2 reversion as it is transmitted in an epidemiological model previously calibrated during an mOPV2 clinical trial performed in Matlab, Bangladesh (20). The combined model allows us to assess the consequences of vaccine reversion in the context of an actual epidemiological setting. By unifying evolutionary theory, population genetics, and epidemiology into a single framework, our study provides a holistic understanding of cVDPV2 emergence as it relates to both genetic reversion and transmission.

## Results

### Brief model structure overview

We previously developed agent-based multiscale epidemiology model designed to replicate household community structure and Sabin 2 vaccine transmission during an mOPV2 clinical trial performed in Matlab, Bangladesh (20, 21). Transmission is contact based and made heterogeneously depending on household community structure membership (**Figure 1A**). Susceptibility is defined by a dose-response model that determines the probability of infection given the strain-specific viral infectiousness, the total viral exposure dose per contact, and the immunity of the recipient host. The total viral exposure dose is determined by the viral shedding concentration of the infected individual and the average fecal-oral dose per contact in the population. Immunity is defined by the OPV-equivalent antibody titer that determines infection probability. Individuals with low immunity upon infection have longer shedding durations and higher viral shedding titers. Further details of the epidemiology model are described elsewhere (21, 22).

**Figure 1.**
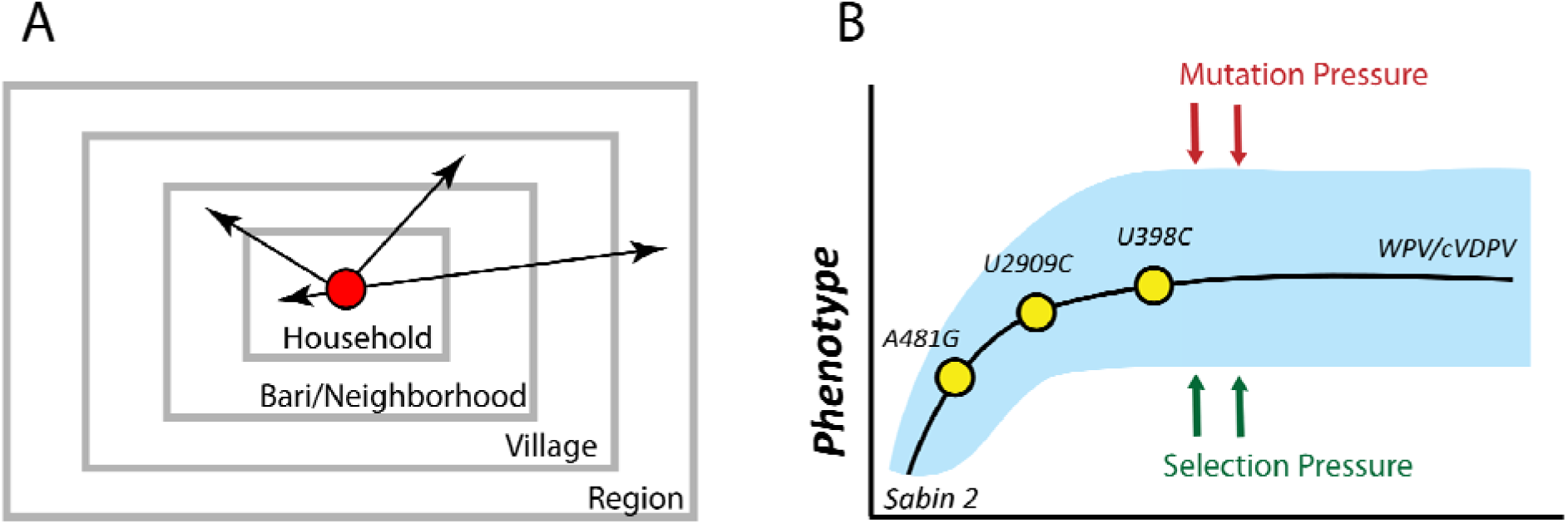
Sabin 2 transmission and reversion schematic. **A)** Multiscale epidemiology model. Individuals are organized into a series of nested, demographic scales defined by households, *baris*/neighborhoods, and villages that together define the greater region. The *bari* is an intergenerational living arrangement of closely related individuals specific to Matlab, the study population the epidemiology model was calibrated to. Conceptually, it is analogous to a neighborhood. Infectious contacts occur at different rates to individuals in each of these demographic scales. During each infectious contact, infected individuals transmit a viral dose that depends on their individual viral shedding concentration and the average fecal oral dose per contact in the population. **B)** Sabin 2 reversion model. Sabin 2 evolution is modeled as a population of competing viral lineages whose average phenotypic end state distribution is identical to that of WPV. Reversion is driven by the acquisition of three gatekeeper mutations (A481G, U2909C, U398C) and the acquisition of deleterious mutations introduced through mutation and purged through selection.

Here, we developed and integrated a new Sabin 2 reversion model into the multiscale epidemiology model. Sabin 2 evolution is modeled as a population of multiple viral lineages founded by a Sabin 2 vaccine virus strain during vaccination. These viral lineages evolve independently and compete for the limited availability of naïve or low-immunity individuals in the population. Phenotypic evolution is driven by the reversion of three gatekeeper mutations (A481G, U2909C, and U398C) and the accumulation of deleterious nonsynonymous mutations generated through mutation and purged through selection or genetic drift (**Figure 1B**). We assume complete genetic reversion results in a viral population whose average infectiousness and shedding duration is identical to those of WPV. Due to the independent acquisition of mutations across different viruses and viral lineages, individual viruses have different infectiousness and shedding duration phenotypes even after the three gatekeeper mutations have reverted.

### Simulating Sabin 2 molecular evolution

To inform molecular evolution, intra-host substitution rates for the three gatekeeper mutations were based on estimates previously calculated from 241 Vp1 (viral protein 1) segments from Sabin-like poliovirus collected during routine surveillance in Nigeria (10, 23). Nonsynonymous and synonymous substitution rates were calibrated (**SI Appendix**) to a set of whole genome sequences and VP1 capsids segments isolated from infections from confirmed Sabin 2, Sabin-like, vaccine-derived poliovirus (VDPV) and cVDPV2 cases (**Figure 2**). One notable feature of poliovirus evolution is the discordance between short-term evolution following vaccination and long-term evolution after prolonged circulation. Short-term evolution is dominated by the accumulation of nonsynonymous mutations, as evidenced by the increased rate of nonsynonymous mutation accumulation in the whole genome sequences during the first year of evolution (**Figure 2B**), the reduction in the ratio of nonsynonymous to synonymous mutations over time (**Figure 2D**), and the elevated dN/dS ratios of Sabin-like samples compared to cVDPV2 samples (Supplemental **Figure 1**).

**Figure 2.**
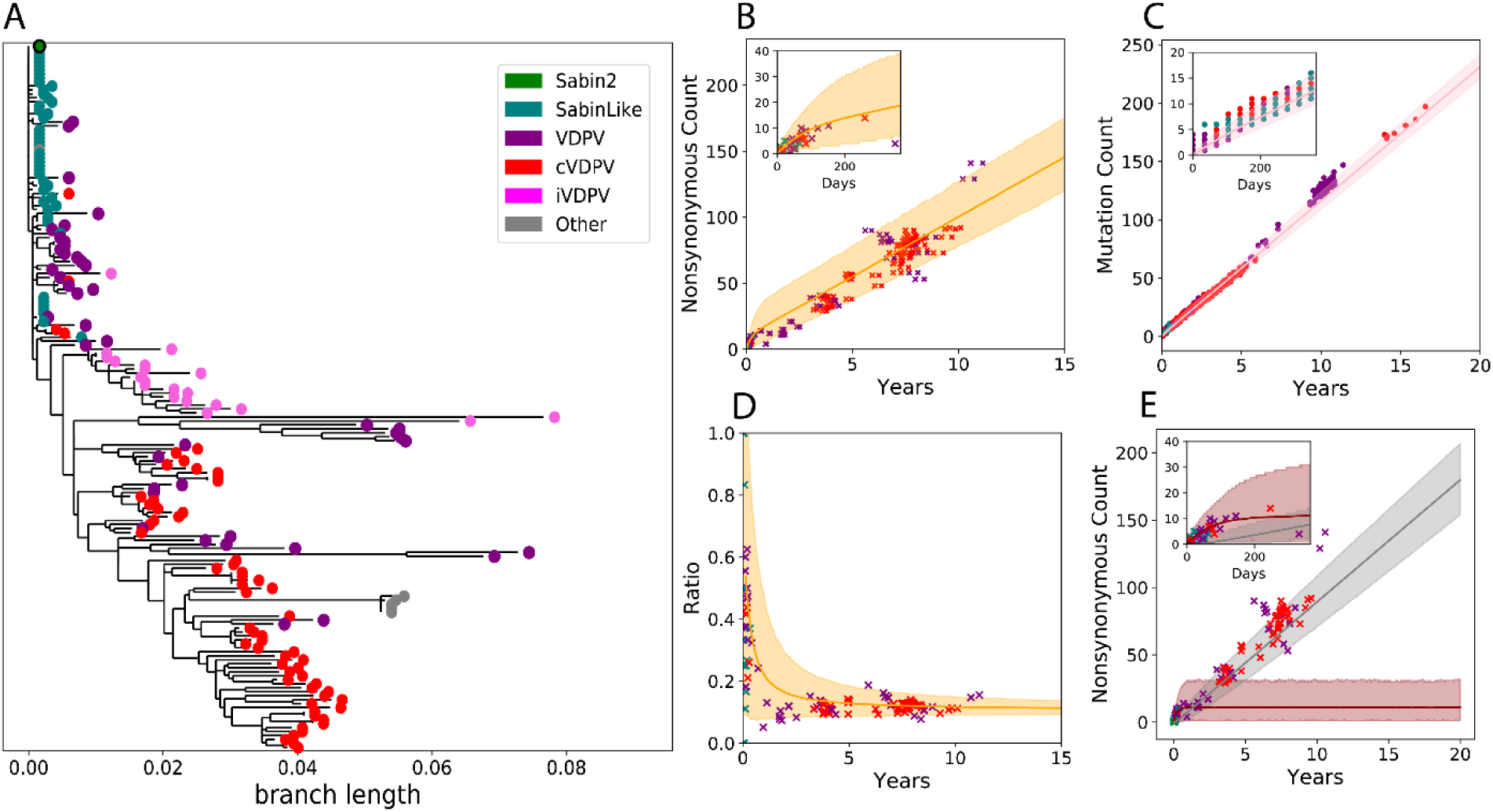
Molecular evolution. **A)** Phylogenetic tree based on the nonsynonymous mutations from the 177 whole genome sequences used to calibrate our model. Samples were categorized as Sabin 2, Sabin-Like, vaccine-derived poliovirus isolated from immunocompromised individuals (iVDPV), vaccine-derived poliovirus (VDPV), and cVDPV2. The other category includes the Lansing and MEF-1 laboratory strains as well as several engineered strains derived from the Lansing strain. Samples classified as iVDPV or “other” were excluded from model calibration. **B)** Simulated genome-wide nonsynonymous mutation accumulation over time (*orange*) compared against those observed in the Sabin 2, Sabin-Like, VDPV, and cVDPV2 whole genome sequences. **C)** Simulated (*pink*) VP1 mutation (synonymous and nonsynonymous) accumulation over time compared against the mutation counts observed in 1,643 VP1 segments. **D)** The ratio of nonsynonymous and synonymous mutations throughout the genome over time. E) Simulated genome-wide nonsynonymous counts partitioned into deleterious (*brown*) and neutral (*grey*). Across all figures, the solid line is the mean simulation outcome and the shaded area marks the boundaries of the middle 95%. Empirical data is represented by points and colored according to the legend in panel A. Time was calculated by dividing the number of synonymous mutations per sample by the synonymous substitution rate (3.16E-05 substitutions/bp/day). The insets in **B**,**C**, and **E** are close ups of the first year of evolution to better show short-term evolution dynamics.

We hypothesized that the difference between short-term and long-term evolution represents a shift in the distribution of fitness effects for nonsynonymous mutations. Simulated nonsynonymous mutations were divided into two categories: deleterious mutations that can cause a reduction in either infectiousness or shedding duration, and neutral mutations that have no phenotypic effect. Each category is associated with a different substitution rate and deleterious nonsynonymous mutations are purged using a recombination-like process that mimics selection and intra-species recombination within the host (**SI Appendix**). Deleterious nonsynonymous mutations dominate molecular evolution during the first year of evolution while neutral nonsynonymous mutations dominate molecular evolution after prolonged circulation (**Figure 2E**).

### Simulating Sabin 2 reversion

Genotype-to-phenotype maps for shedding duration were generated by comparing the shedding duration of naïve individuals vaccinated with Sabin 2 (median 30.3, average 36 days) with those infected with WPV (median 40.3, average 48 days) (**SI Appendix**). Due to the rapid intra-host fixation of the three gatekeeper mutations (**Figure 3A**), we assert that reversion during an infection extends its shedding duration (**Figure 3B**). Prior to any intra-host evolution, the genotype-to-phenotype map for shedding duration estimates that naïve individuals vaccinated with non-reverted Sabin 2 have a median shedding duration of 13.5 days (**Figure 3B**, *dashed red line*). The clinically-observed shedding durations of Sabin 2 vaccinated individuals (**Figure 3B**, *solid red line*) is realized as the average shedding durations of different viral genotypes composed of unmutated and reverted Sabin 2 viruses (**Supplemental Table 1**). After vaccination, 7.6% of simulated vaccinated individuals shed non-reverted Sabin 2 at the end of shedding, 92.4% shed virus with at least one reverted gatekeeper mutation, and 35.2% shed virus with all three gatekeeper mutations reverted. Interestingly, we did not identify a deleterious cost (reduction in shedding duration) associated with nonsynonymous mutation accumulation (**SI Appendix**).

**Figure 3.**
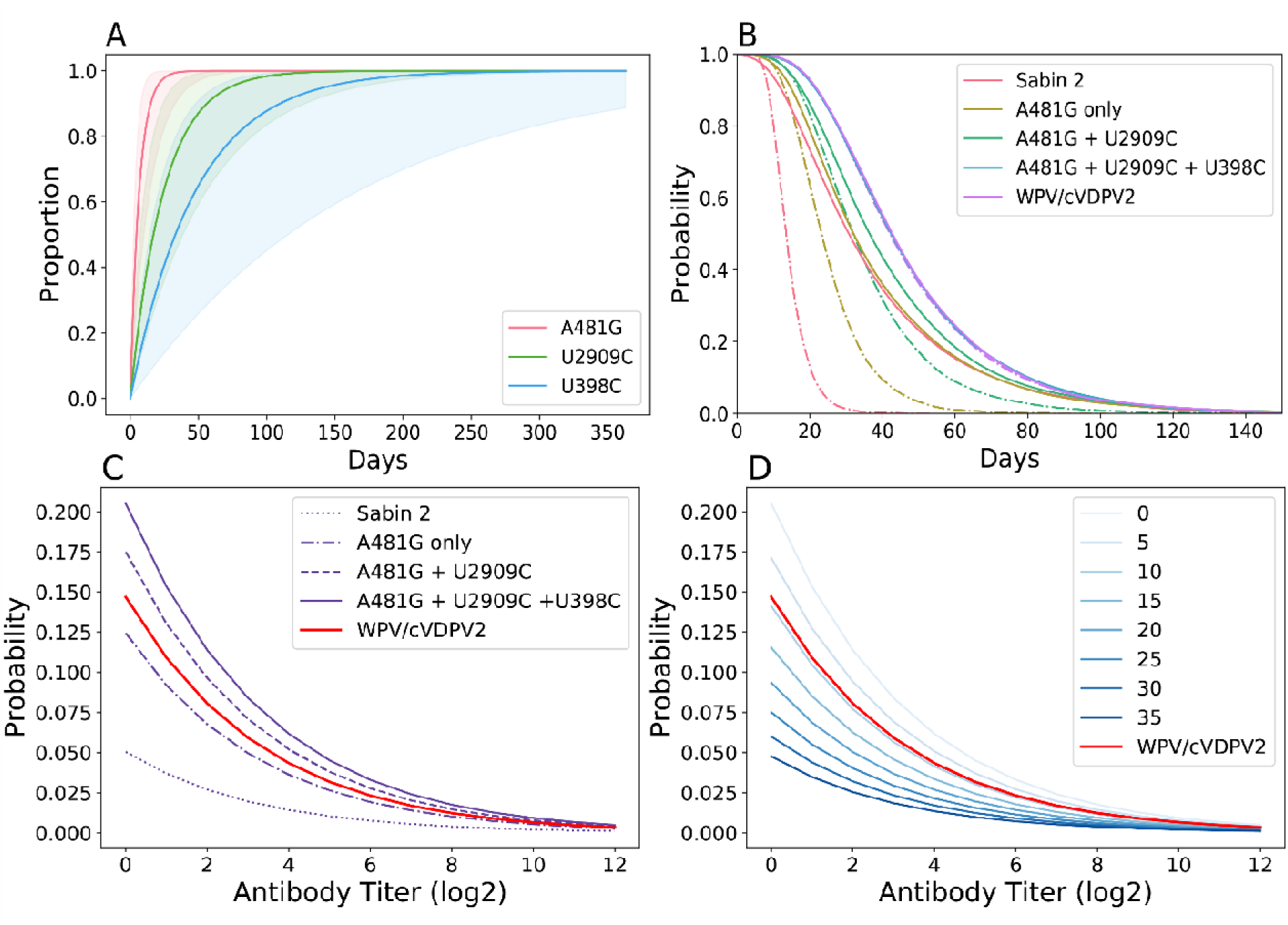
Phenotypic Evolution. **A)** Cumulative density reversion probability functions for the three gatekeeper mutations. The solid line is the mean and the shading the 95% confidence interval. **B)** Shedding duration profile of immunologically naïve individuals infected with Sabin 2, WPV/cVDPV2, and three different intermediate viral genotypes. The dotted lines are the shedding durations of the initial viral genotype assuming no further reversion or evolution. The solid line are the shedding durations of each viral genotype where the reversion of new gatekeeper mutations extends the current shedding duration. Nonsynonymous mutations have no discernable phenotypic effect with regards to shedding duration. **C and D)** Viral infectiousness. Each curve shows strain-specific probability of infection given a single CCID50 dose plotted against immunity. Panel **C** shows infectiousness following the acquisition of each of the three gatekeeper mutations. Panel **D** emphasizes the role of deleterious nonsynonymous mutation and shows the infection probability of different viral genotypes with all gatekeeper mutations and deleterious nonsynonymous mutations count ranging from zero to 35.

The genotype-to-phenotype map describing viral infectiousness was based on clinical differences of human dose response between Sabin 2 and WPV. Unlike shedding duration, we identified a small negative effect associated with deleterious nonsynonymous mutations resulting in a reduction in viral infectiousness (**SI Appendix**). Our model predicts that viral genotypes with no deleterious nonsynonymous mutations but are reverted at A481G and U2909C are more infectious than typical WPV strains (**Figure 3C**). Each additional deleterious nonsynonymous mutation reduces infectiousness such that a viral genotype with all three gatekeeper mutations reverted and nine deleterious mutations is as infectious as WPV (**Figure 3D**).

### Examining the evolutionary epidemiology dynamics of Sabin 2 reversion

To explore how Sabin 2 reversion and transmission interact, we simulated transmission following an mOPV2 campaign targeting 10% of children under five in a population with 10 times the fecal-oral dose of Matlab (2.5e-6 grams per contact) five years after vaccination cessation (**Figure 4**). These epidemics are cyclic (**Figure 4A**) and characterized by boom-bust cycles that eventually stabilize within the first three years of transmission. Under these conditions, 10% of stochastic replicates failed to establish stable, endemic transmission. These stochastic replicates failed after the initial expansion wave collapsed (**Supplemental Figure 2A**). Should transmission recover, the epidemic rebounds and the proportion of individuals with no immunity (serum neutralizing antibody titer < 8) declines while the proportion of individuals with high immunity (serum neutralizing antibody titer > 256) increases (**Figure 4B**). Towards the end of the initial epidemic, the proportion of individuals with no immunity begin to increase due to a combination of new births and the waning of pre-existing immunity and sets up the conditions for the next wave of infections.

**Figure 4.**
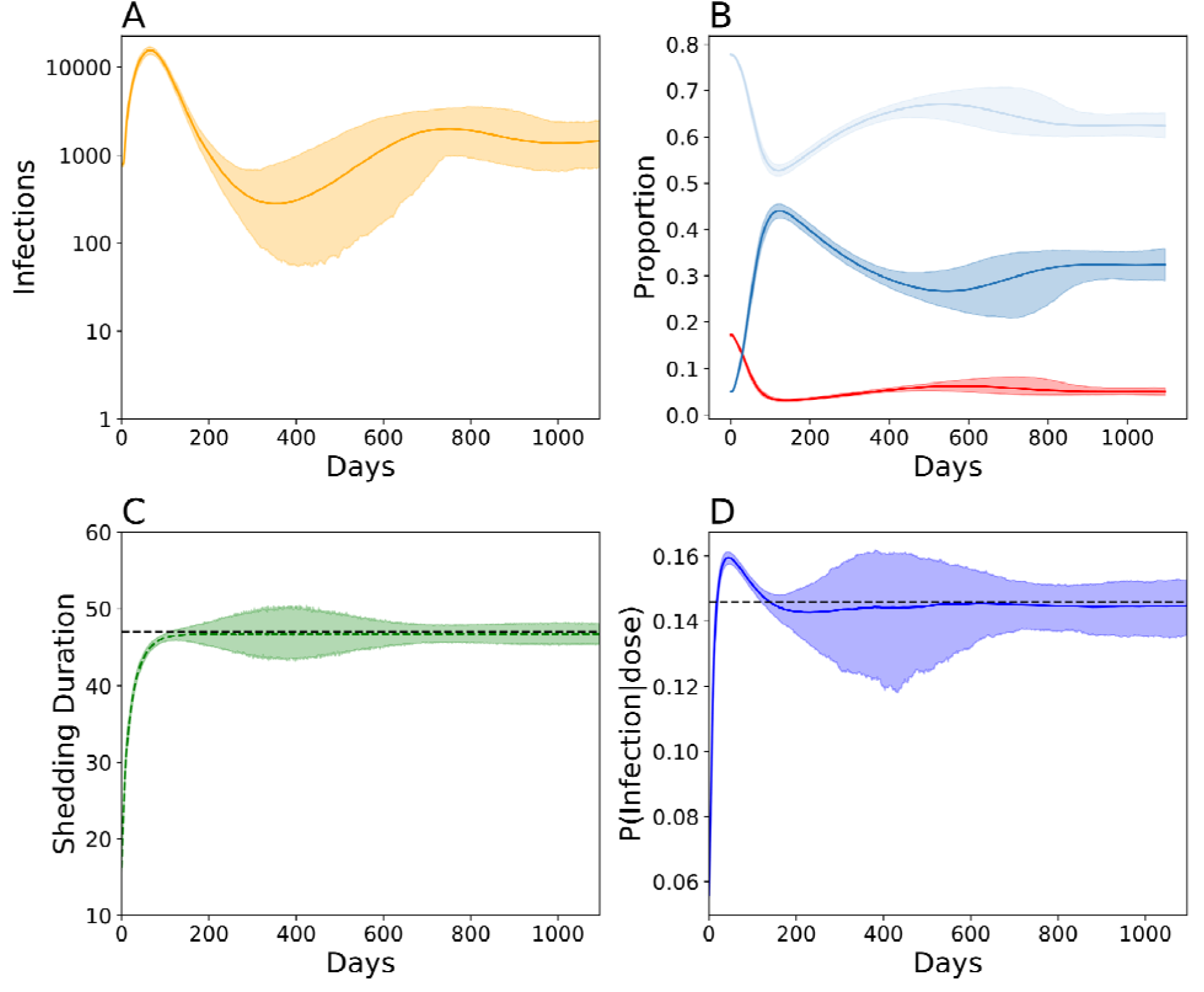
Evo-epidemiological dynamics following a mass vaccination campaign targeting 10% of children under five, five years post-vaccination cessation in Matlab, Bangladesh. Simulations assume 10x the fecal oral dose in Matlab to ensure stable transmission and conditioned on stable, endemic transmission for at least three years. **A)** Total number of infected individuals over time. Solid line indicates the mean and the shading the boundaries of the middle 95% from 200 simulations **B)** Longitudinal immune profiles for individuals with low immunity (*red*, antibody titer < 8), intermediate immunity (*light blue*, antibody titer > 8 and < 256), and high immunity (*dark blue*, antibody titer > 256). **C)** Shedding duration evolution in naïve individuals over time. D) Viral infectiousness evolution over time. For panels B-D, the solid line denotes the simulated average and the boundaries the 95% confidence interval around the mean. For panels **C** and **D**, the dashed black line indicates the average WPV phenotype.

Complete Sabin 2 reversion (**Figure 4C-D**) occurred within the first 200 days of transmission. To describe the evolutionary dynamics of the simulated viral population, we focused on the expected mean (the average of the average) phenotype and the variance of the expected mean. While both shedding duration and infectiousness converged to the WPV phenotype, shedding duration approached it monotonically while infectiousness exceeded WPV during the first few months of transmission. The expected means of both phenotypes were not sensitive to changes in infection number. However, the variance of the expected mean fluctuated over time and was sensitive to epidemiological dynamics. This variance was smallest at the onset of the epidemic when infection numbers were highest and biggest around day 400 when infection number was the smallest. The variance of the expected mean was inversely correlated with total infection number, but its magnitude also depended on the rate with which the epidemic was increasing or decreasing over time (**Figure 4, Supplemental Figure 3**).

Surprisingly, complete Sabin 2 reversion does not guarantee stable transmission. The simulations with observed transmission failure did not show a lack of evolutionary fitness: the simulated viral populations in these simulations had infectiousness and shedding durations equivalent to or exceeding that of WPV (**Supplemental Figure 2B-C**).

### Population-level immunity and hygiene are major constraints of cVDPV2 emergence

We next utilized this model to simulate cVDPV2 emergence and understand risk. Our scenarios included a point importation of an infant vaccinated with Sabin 2, and a mass vaccination campaign with up to 80% coverage in children under five years of age. We explored scenarios with fecal oral doses equal to, five times greater, and 10 times greater than Matlab (**Figure 5**). The fecal oral dose, combined with the infection-specific, time-dependent viral shedding concentration per gram of stool (22), determines the total exposure dose per infectious contact. cVDPV2 emergence was defined as an outbreak significant enough to trigger a warning from passive routine surveillance efforts. However, because surveillance is not currently simulated, we operationally define cVDPV2 emergence as a simulation with sustained cVDPV2 transmission for at least three years.

**Figure 5.**
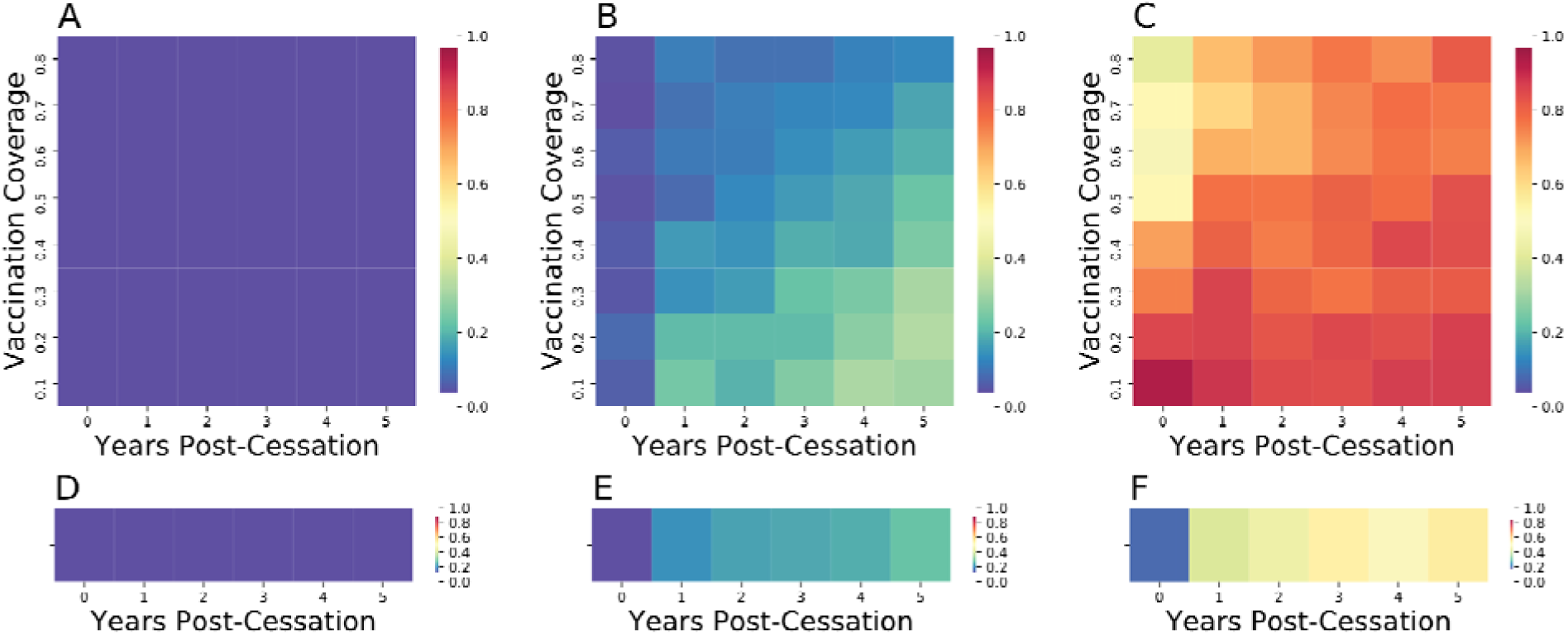
cVDPV2 outbreak risk. cVDPV2 emergence after a mOPV2 vaccination campaign **(A-C)** or importation of a single infant immunized with Sabin 2 **(D-F)** occurring after up to five years of vaccination cessation with fecal-oral doses equivalent to **(A/D)**, five times greater **(B/E)**, and 10 times greater **(C/F)** than that inferred for Matlab during an mOPV2 clinical trial.

We found zero risk of cVDPV2 emergence following a mass vaccination campaign (**Figure 5A**) or point importation event (**Figure 5D**) in Matlab with its current fecal oral dose. However, cVDPV2 emergence is possible at higher fecal-oral doses. Under these conditions, cVDPV2 emergence risk increases with time since vaccination, but is controlled within the community by higher vaccination coverage. When the fecal-oral dose was five times that of Matlab (1.8e-6 grams per contact), all vaccination campaigns and point importations immediately after vaccination cessation (year 0) had a less than 0.05 chance of causing a cVDPV2 emergence (**Figure 5B**). Vaccination campaigns with less than 30% coverage in populations with more than three years of vaccination cessation had the highest risk of cVDPV2 emergence, with an emergence rate between 20% and 30%. Following point importation, cVDPV2 emergence risk rises from 0% immediately after cessation to 20% after five years of cessation (**Figure 5E**).

When the fecal-oral dose was ten times that of Matlab (2.5e-6 grams per contact), high vaccination coverage could not efficiently prevent cVDPV2 emergence (**Figure 5C**). Immediately after vaccination cessation, simulations with high vaccination coverage (> 70% coverage) resulted in cVDPV2 emergence 40-50% of the time. All other scenarios had a greater than 60% chance of cVDPV2 emergence. This increases to a greater than 80% chance following a vaccination campaign with less than 30% coverage in populations with more than three years of vaccination cessation. cVDPV2 emergence risk following a point importation event increases sharply with time since vaccination (**Figure 5F**), increasing from 5% immediately after cessation to 55% after five years of cessation.

## Discussion

cVDPV2 outbreaks are serious public health dilemmas made possible by viral evolution and exacerbated by epidemiological conditions. By unifying evolutionary and epidemiological theory into a single framework, our model assesses cVDPV2 emergence risk in relation to changes in shedding duration and infectiousness associated with vaccine reversion and the constraints to transmission imposed by immunity, sanitation, and contact structure. To overcome the lack of direct transmission measurements for Sabin 2 reversion intermediates, we generated genotype-to-phenotype maps for shedding duration and viral infectiousness by relying on established molecular evolution and population genetic theory.

Sabin 2 reversion can be viewed as an adaptive process where Sabin 2 must climb up a fitness landscape to achieve the optimal, WPV-like phenotype. Within the poliovirus virology literature, Sabin 2 is commonly presented as a purely adaptive process where fixed mutations contribute positively to the emergence of the WPV phenotype (24). However, it is unlikely that all fixed nonsynonymous mutations are beneficial (25) and viruses must purge deleterious mutations to prevent Muller’s Ratchet from causing an irreversible decline in viral fitness (26, 27). Muller’s ratchet can be avoided by engaging in intra-species or inter-species recombination with Enterovirus C viruses (28) and by altering the distribution of fitness effects (DOFE) for fixed mutations (29, 30). The latter is based on Fisher’s Geometric Model (FGM) of adaptive evolution, which predicts that mutations with large-phenotypic effects are unlikely to fixate the closer an organism is to its fitness optimum. Our model incorporates FGM by attributing large phenotypic effects to the gatekeeper mutations and specifying nonsynonymous mutation rates so that early adaptive evolution is dominated by deleterious nonsynonymous mutations while long-term evolution is dominated by neutral ones.

Interestingly, we failed to identify a deleterious cost associated with nonsynonymous mutations and shedding duration but did identify one associated with nonsynonymous mutations and infectivity. This finding suggests that there are differences between intra-host and inter-host selection. This could reflect strong purifying selection induced by the highly competitive intra-host survival dynamics due to high viral titers. Conversely, the deleterious cost associated with infectiousness could result from the genetic drift induced by the severe sampling of viral genotypes during person-to-person transmission (25, 31). Strong genetic drift could weaken the efficacy of purifying selection in removing deleterious mutations associated with inter-host transmission, but more work is needed to assess differences in inter- and intra-host selection. The phenotypes we modeled are the consolidation of multiple factors such as cell invasion, replication, immune evasion, and environmental survival. These subfactors likely contribute to both shedding duration and infectiousness, but our ability to map molecular evolution to these subfactors remains limited.

At its core, cVDPV2 emergence is an evolutionary and epidemiological problem that requires a combined approach to properly assess how the two interact. From an evolutionary perspective, the availability of susceptible hosts and the distribution of transmissions dictates the strength of genetic drift. Once the gatekeeper mutations are fixed, the phenotypic stability of the viral population is maintained as a mutation-selection-drift balance and the range of phenotypic values explored by the viral population is determined by the total number of infected individuals in the population. From an epidemiological perspective, Sabin 2 reversion causes infectiousness and shedding duration to rapidly increase within the first 200 days of vaccination and allow more individuals to become infected. Despite the emergence of WPV-like viruses in our simulations, complete genetic reversion of Sabin 2 does not guarantee a cVDPV2 outbreak. Whether these viruses cause a cVDPV2 outbreak requiring public health intervention depends on population-level immunity and the average viral exposure dose per infectious contact (4, 6, 22, 32).

The current strategies for controlling cVDPV2 involve implementing aggressive mOPV2 SIA activities in outbreak response zones and neighboring regions, intensifying routine immunization in high-risk areas with inactivated polio vaccine (IPV), ensuring a sufficient supply of Sabin 2 OPV for future eradication strategies, and identifying high cVDPV2 outbreak risk zones (33). Our work clearly shows that even regions with historically high vaccination rates are prone to cVDPV2 outbreaks if sanitation is poor. Evaluations of high cVDPV2 outbreak risk regions should include sanitation assessments and utilize tools designed to measure fecal contamination for other fecal-oral pathogens such as *E. coli* (34). Prior to mass immunization with OPV2, sanitation assessments should be made to assess the risk of a cVDPV2 outbreak in both the targeted intervention zone and in the surrounding regions. These assessments will be useful for resource allocation and for determining the minimum coverage needed to prevent future cVDPV2 outbreaks. Regions with poor sanitation will require aggressive, high-coverage SIA vaccination campaigns to prevent seeding future cVDPV2 outbreaks.

Sanitation assessments will also help public health officials determine whether additional WASH (water, sanitation, and hygiene) interventions are needed to supplement ongoing vaccination activity. Our study reconfirms that there are sanitation regimes defined by high fecal oral doses that is guaranteed to seed future cVDPV2 outbreaks (22), regardless of vaccination coverage. Although long-term investments in infrastructure are prohibitively expensive, relatively simple measures such as the installation of sanitary latrines and drinking water tubewells have been effective at reducing fecal contamination in Matlab, Bangladesh (35). Optimistically, short-term sanitation interventions may be effective if implemented similarly to an emergency response campaign (36), especially if they are targeted at vaccine recipients and their immediate household community members (21).These targeted, short-term sanitation interventions could alleviate some of the challenges associated with non-compliance, long-term adherence failure, and poor implementation that reduce the efficacy of standard WASH interventions (37, 38).

Our model lays the foundations for integrating population genetic theory and epidemiology to study the evolutionary epidemiology dynamics of rapidly evolving infectious diseases. The success of our framework hinges on an understanding of a pathogen’s evolutionary trajectory as well as the initial and ending phenotypic states. We anticipate our model will be extended to the novel OPV2 (nOPV2) candidates (39) as more about their clinical phenotypes and molecular evolution are known. Depending on how attenuated and evolutionarily stable the novel OPV2 candidates are relative to Sabin 2, mass nOPV2 vaccination may reduce the need to also control sanitation levels. We also anticipate our model will be useful for assessing the risks associated with the live-attenuated varicella zoster vaccines, which also shows strong evidence of attenuation reversion conferred by a commonly accessed evolutionary pathway (40).

## Materials and Methods

### Sequencing Data

We downloaded fasta files for the VP1 segments and whole genome sequences of Sabin 2 and Sabin 2 derived genomes from Genbank. These samples were collected across 102 studies performed in countries including Egypt, Madagascar, China, Nigeria, the United States, Israel, Switzerland, and the Democratic Republic of Congo. Multiple sequence alignments were first generated using MUSCLE (v3.8.31) (41). Sequences were compared to a known Sabin 2 sample (accession ID: AY184220) to identify and remove insertions. Sequences with long internal gaps (>15 bases) and coverage < 30% (percentage of called bases) were discarded. Coding sequence alignments from the whole genome sequences were generated for each of the poliovirus coding regions by generating multiple sequence alignments for each of the 15 translated protein coding sequences of the poliovirus genome. These alignments were further filtered to remove samples with < 90% coverage.

A total of 177 whole genome sequences and 1,643 VP1 sequences were retained for analysis. Samples were classified as Sabin 2, Sabin-Like, VDPV (vaccine-derived poliovirus), iVDPV (immunodeficiency-related vaccine-derived poliovirus), cVDPV2 based on their original study classifications. Viruses that displayed <1% VP1 nucleotide sequence difference from the parental Sabin 2 genome were classified as Sabin-lik2 and those with 1-15% nucleotide sequence difference were classified as VDPV. VDVPVs isolated from immunodeficient individuals were classified as iVDPV while samples with evidence of having originated from prior transmission are classified as cVDPV2. The whole genome sample set also included the Lansing and MEF-1 laboratory strains as well as several engineered Lansing strain derivatives. Accession numbers, classifications, and study origin for each sample are in **Supplemental File 1** and **Supplemental File 2**. FASTA files for the aligned VP1 and whole genome coding sequences are available on the **Github repository**.

Coding sequence alignments were used to generate a phylogenetic tree using RAxML (v8.2.10) with a general time reversable CAT (GTRCAT) model (42). Estimates of nonsynonymous and synonymous mutation counts and dN/dS were based off the RAxML generated phylogenetic tree. Estimates of nonsynonymous and synonymous mutation counts from the VP1 sequences were counted directly from the sequence data. Nonsynonymous and synonymous mutation counts for codons with multiple mutations were averaged across all possible, unweighted evolutionary paths.

## Model Description

### Defining Genotypes

Genotypes are defined relative to the Sabin-2 genome and defined by the reversion state of three gatekeeper mutations (A481G, U2909C, and U398C), and the number of newly derived nonsynonymous and synonymous mutations in the rest of the genome. The Sabin-2 genome is defined as an array with 6 different mutation classes:

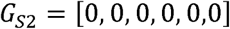

The first three are Boolean flags that indicate the allelic states of A481G, U2909C, and U398C (0 = unreverted, 1 = reverted). The last three elements represent the total number of deleterious nonsynonymous, neutral nonsynonymous, and synonymous mutations present in the genome. The counts in the latter categories exclude the three gatekeeper mutations. Splitting nonsynonymous mutations into neutral and deleterious simplifies the true distribution of fitness effects and was necessary to reconcile phenotypic and molecular evolution (**SI Appendix**). These genotypes represent the consensus sequence of fixed variants within vaccinated and infected individuals.

For evolved poliovirus strains, we use the notation:

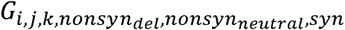

where *i, j*, and *k* are replaced with the allelic states of A481G, U209C, and U398C. *nonsyn*_*neutral*_, *nonsyn*_*del*_, and are replaced with their respective counts or replaced with X if these mutation classes are irrelevant. For example, *G*_0,0,0,0,0,0,0_ is equivalent to the unmutated Sabin-2 vaccine strain and (*G*_*S*2_) *G*_1,1,1,10,2,15_ is a genotype where all three gatekeeper mutations have reverted and where 10 deleterious nonsynonymous, 2 neutral nonsynonymous, and 15 synonymous mutations have accumulated.

### Genomic evolution: Modeling Sabin 2 molecular evolution

Genomic evolution is modeled using intra-host substitution rates (λ), the fixation of newly derived mutations within the host, for each mutation class (*λ*_*A481G*_, *λ*_*U*2*909C*_, *λ*_*U3*98*C*_, *λ*_*syn*_, *λ*_*nonsyn,del*_, *λ*_*nonsyn,neutral*_). This substitution rate reflects the true de novo mutation rate, genetic drift, and selection within the host. This simplification allows us to use a single consensus genome to represent the fixed mutations present in the intra-host viral population. Each mutation class (the three gatekeeper mutations, synonymous mutations, deleterious nonsynonymous mutations, and neutral nonsynonymous mutations) is associated with a substitution rate that describes its expected fixation time (**Supplemental File 3**).

To accommodate the effects of intra-host competition and recombination, deleterious nonsynonymous mutation accumulation also depends on a recombination-like mechanism with rate *λ*_*recombination*_ that purges deleterious mutations from the genome (26, 28, 43). Intra-host substitution rates for the gatekeeper mutations were used as described in (10) while, *λ*_*syn*_, *λ*_*nonsyn,del*_, *λ*_*nonsyn,neutral*_, *λ*_*neutral*_, and *λ*_*recombination*_ were calibrated to the whole genome sequences and VP1 segments downloaded from Genbank. Only confirmed Sabin 2, Sabin-Like, VDPV, and cVDPV samples were used for calibration. All other samples were excluded from calibration. Calibration details are provided in the **SI Appendix**.

Upon vaccination with Sabin 2, fixation times for all mutation classes are calculated by drawing from exponential distributions whose rate parameters are defined by the substitution rate of each mutation class. For the three gatekeeper mutations, these fixation times determine the time to reversion. For nonsynonymous and synonymous mutations, these fixation times determine the time to the next substitution event. To prevent Mueller’s Ratchet from resulting in infinite deleterious mutation load, we also draw a time to the next “recombination” event from an exponential distribution defined *λ*_*recombination*_ by. During each “recombination” event, the model draws a random number from a discrete, uniform distribution ranging from zero to *n*_*non syn,deleterious*_, the number of deleterious nonsynonymous mutations in the genome, to determine the number of deleterious mutations to remove. Conceptually, this represents purifying selection within the host and allows us to accommodate the assumed increase in purifying selection as Sabin 2 reverts to cVDPV2. Pseudocode is provided in ***Supplemental Figure 4***.

### Genotype to Phenotype: Mapping molecular genetics to shedding duration and infectiousness

A core assumption of our model is that each of the three gatekeeper mutations contributes to the final, WPV phenotype proportional to their relative substitution rates. This assumption is based in part on evolutionary fitness landscape and adaptation theory, which posits that beneficial mutations of large effect are more likely to be fixed early on when an organism is far the optimal fitness peak on a simple, non-rugged fitness landscape (29, 44). By definition, attenuated Sabin 2 vaccine strains are far from the optimal fitness peak for human infection and genetic reversion is the process of climbing the fitness landscape to reach the optimal, WPV-like phenotype. Because of their fast substitution rate, we attribute the bulk of phenotypic change to A481G, followed by U2909C, and to a lesser extent U398C. The ranked ordering of phenotypic effect in our model is supported in the literature. A481G and U2909C are consistently identified as the primary molecular determinants of reversion in *in vitro* and *in vivo* studies (11–16). U398C is only occasionally identified by these studies but its recurrent fixation within the first few months of transmission (9, 10) suggest its adaptive benefit may be too small to be assessed in laboratory-based assays relative to A481G and U2909C.

Two epidemiologically relevant phenotypes were calculated for each infection based on its corresponding genotype: 1) shedding duration and 2) infectiousness. Both shedding duration and infectiousness are based on a previously described poliovirus dose-response model (22). Conceptually, the shedding duration is analogous to intra-host selection and infectiousness is analogous to inter-host selection. Details regarding model calibration are deferred to the **SI Appendix**.

We assume shedding duration follows a log-normal survival distribution with the form:

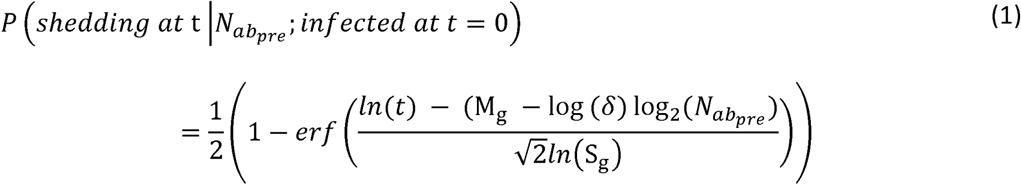

where 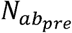 is the pre-infection OPV-equivalent antibody titer (22). For each genotype, M_*g*_ and *S*_*g*_ are genome-specific parameters that influence the average shedding duration and its standard deviation. The mean of this log-normal distribution is defined by 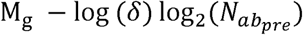 and its standard deviation defined by *S*_*g*_.

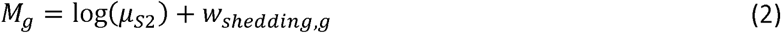

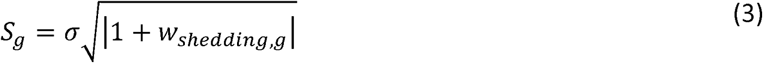

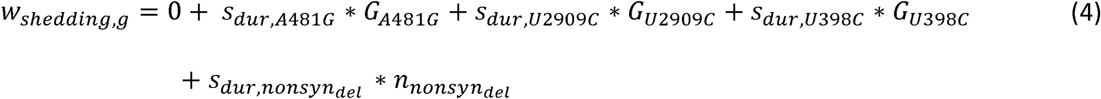

where *μ*_*S2*_ is the average shedding duration of a completely unevolved Sabin 2 virus (*G*_S2_) in immunologically naiive individuals 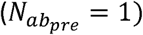, *σ* the standard deviation in shedding duration for *G*_S2_ *w*_*shedding,g*_ defines the shedding duration fitness of a strain, *G*_*A*481*G*_, *G*_*U*2909*C*_, and, *G*_*U*398*C*_ refer to the allelic status of the three gatekeeper mutations, and *S*_*dur,A*481*G*_, *S*_*dur,U*2909*C*_, *S*_*dur,U*398*C*_ are selection coefficients that describe the change in shedding duration conferred by each of these mutations. Note that fitness is additive but in log space.

Upon infection or vaccination, we randomly draw a shedding duration from a lognormal distribution with mean 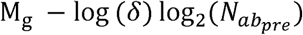 and standard deviation S_g_. A core feature of our shedding duration model is that new gatekeeper mutations arising within the host extends shedding duration. If a new gatekeeper mutation is acquired, the shedding duration is extended:

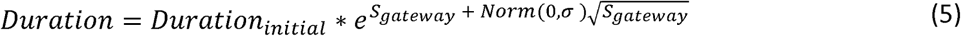

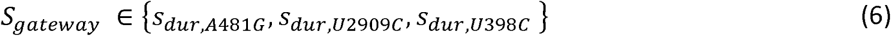

where *D*_*initial*_ is the shedding duration before acquisition of the gatekeeper mutation and, *S*_*gateway*_ is the shedding duration selection coefficient for the gatekeeper mutation. Note that *Duration*_*initial*_ and the increase in shedding duration, 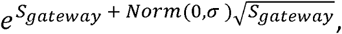, are log-normally distributed and that multiplying two, independent log-normal distributions results in a log-normal distribution whose mean and variance are the sum of the mean and variances of the original distributions. As a result, shedding duration is the summation of multiple, genome-specific shedding durations whose weights are time-dependent and depend on the fixation of A481G, U2909C, U398C and the accumulation of nonsynonymous mutations.

Infectiousness is defined by a beta-Poisson model that assumes a single infectious unit is sufficient to start an infection and that multiple infectious units contribute independently to the probability of infection:

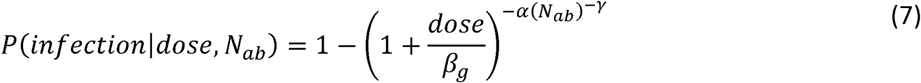

where *α* and *β*_g_ are parameters for a beta-Poisson function, dose is the viral exposure dose, and *γ*, captures the reduction in shedding probability with increasing immunity. Altering the *β* parameter shifts the beta-poisson function along the x-axis (dose) and altering the parameter changes its slope. Differences in infectiousness between Sabin 2 strains and WPV can be modeled using the same *α* but with modified *β* parameters (22). We interpret the *β* parameter as a measurement of strain infectivity and define *β*_*g*_ as:

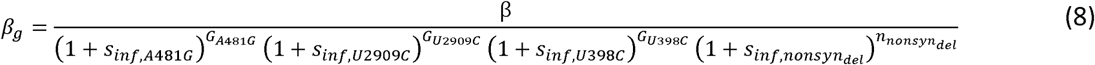

where *β* is the strain infectivity parameter for Sabin 2 (22) (assuming no evolution), 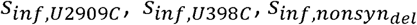 are selection coefficients denoting the change in infectiousness conferred by each of the gatekeeper mutations, and 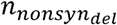 are the number of nonsynonymous mutations in the genome. Parameter estimates were obtained by comparing the CCID50 between Sabin 2 and WPV (**SI Appendix**). The inferred selection coefficients for the three gatekeeper mutations are positive (increases infectiousness) while the selection coefficient for deleterious nonsynonymous mutations was negative (decreases infectiousness). Infectiousness is calculated only at the onset of either vaccination or infection.

## Data Availability

Data Availability
Data and code will be made publicly available at https://github.com/InstituteforDiseaseModeling/MultiscaleModeling/tree/evo_epistasis_upon_publication.

## Acknowledgements

We acknowledge all members of the Institute for Disease Modeling, Global Good, and the Bill and Melinda Gates Foundation for their useful discussion and input. We also acknowledge Adam Lauring and Andrew Valesano for early manuscript edits and comments.

## Funding

Funding was provided by Bill and Melinda Gates through the Global Good Fund. Funders had no role in the study design, data collection, analysis, interpretation, or writing of the manuscript. The corresponding author had full access to all the data in our study and had final responsibility for the decision to submit for publication.

## Data Availability

Data and code will be made publicly available at https://github.com/InstituteforDiseaseModeling/MultiscaleModeling/tree/evo_epistasis upon publication.

## Declaration of Interests

The authors declare no competing interests during the study. WW, JG, and MF were employed by the funders through the Bill and Melinda Gates Foundation after study completion.

---

**Supplementary Figure 1.**
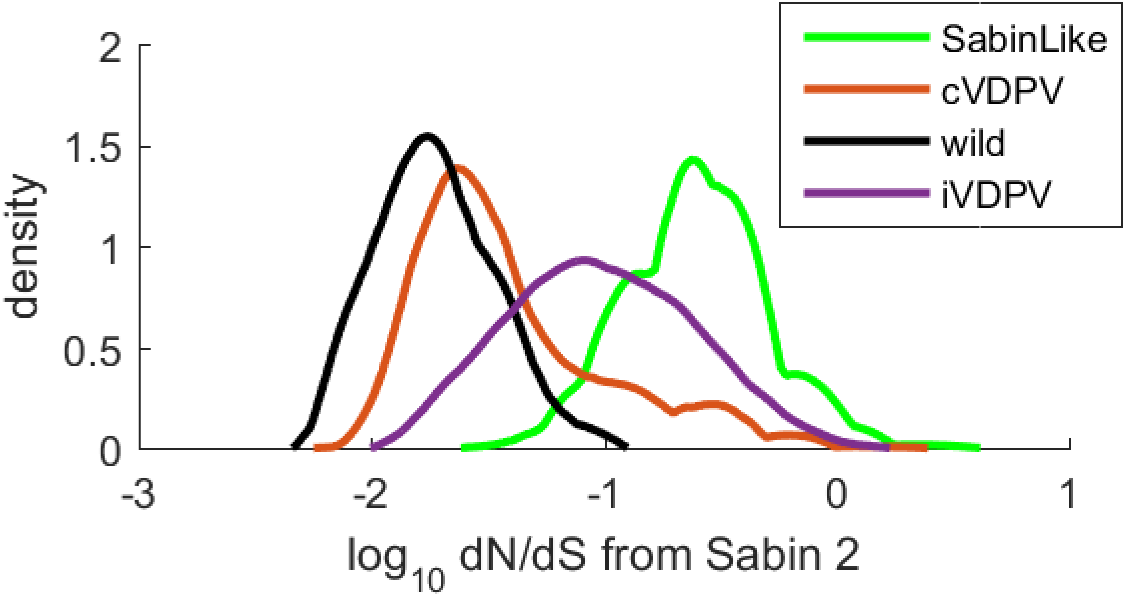
dN/dS ratios for Sabin 2-like, cVDPV2, WPV, and iVDPVs.

**Supplementary Figure 2.**
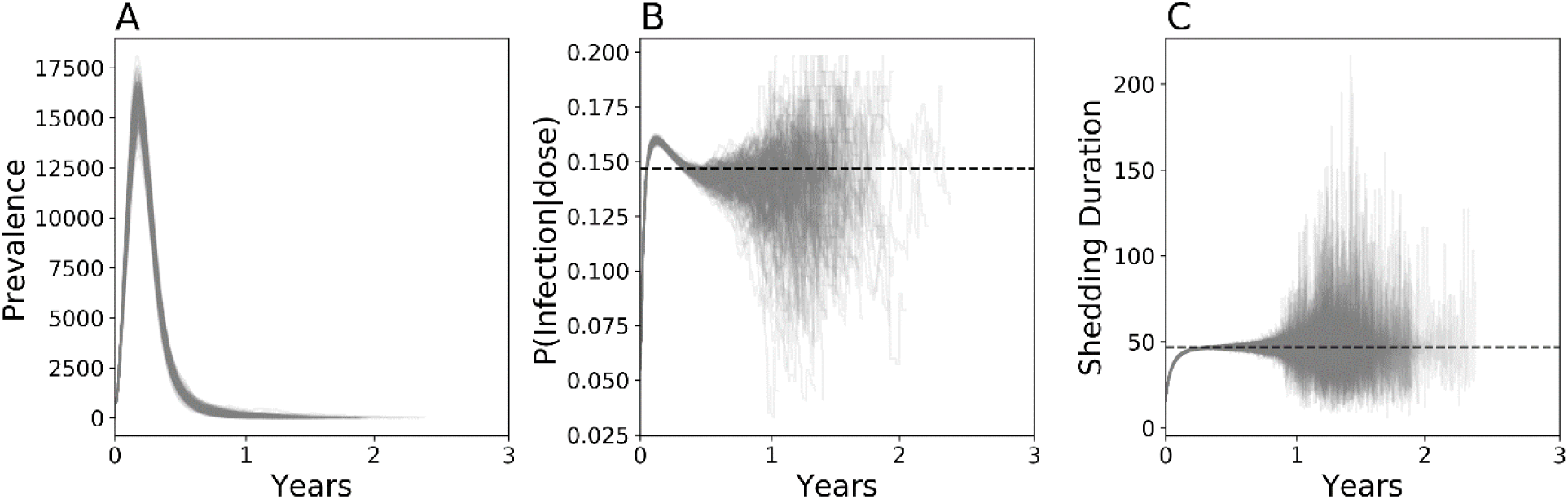
Simulated evolutionary and epidemiology dynamics following a mass vaccination campaign targeting 10% of children under five, five years post-vaccination cessation in Matlab, Bangladesh, conditioned on epidemic extinction within the first three years of transmission. **A)** Total number of infected individuals over time **B)** Infectiousness represented as the probability of infecting an immunologically naïve individual given single CCID50 unit dose. **C)** Shedding duration in immunologically naïve individuals. Each line represents a different simulation outcome.

**Supplementary Figure 3.**
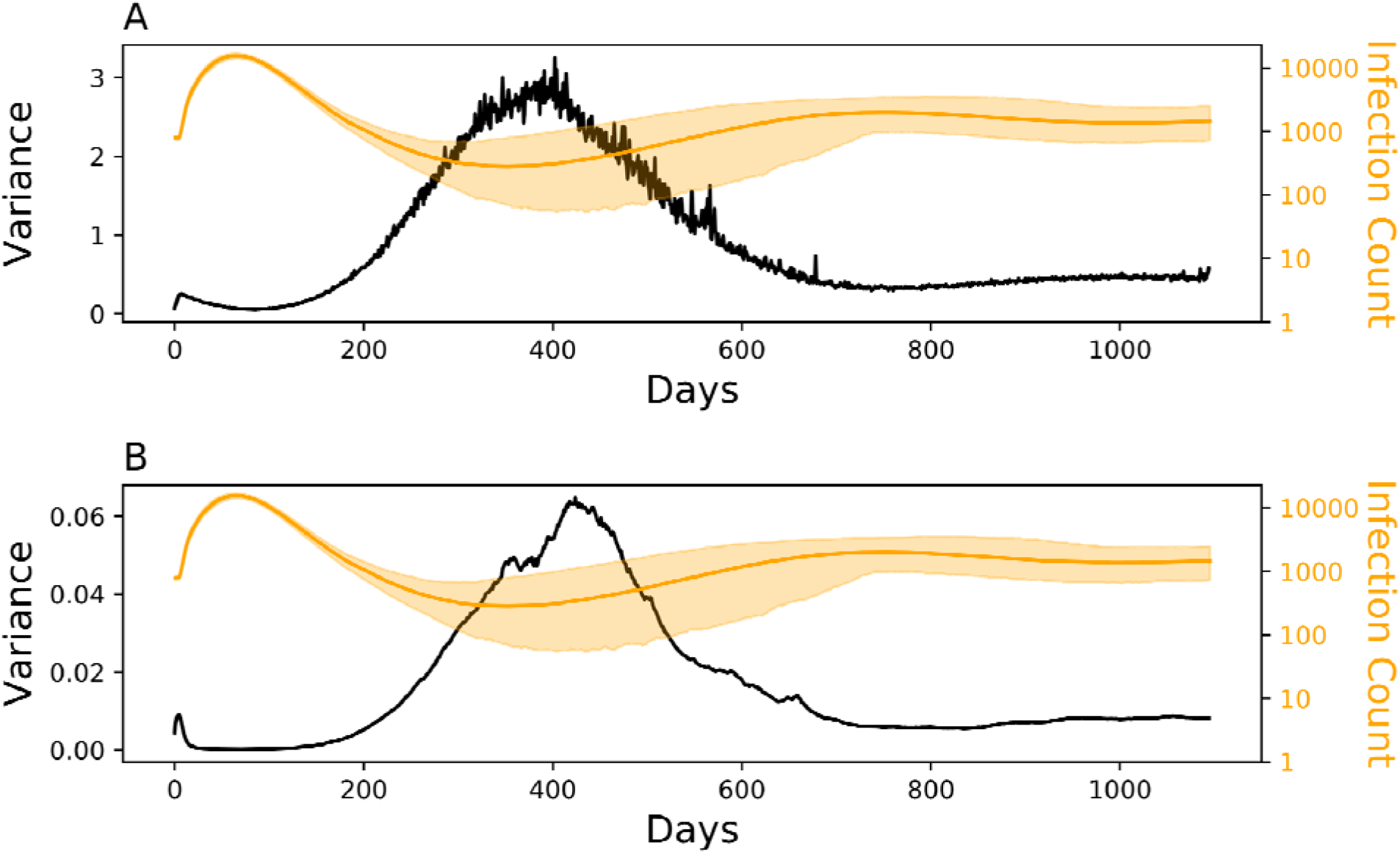
The variance of the expected mean (*black*) for shedding duration **(A)** and infectiousness **(B)** compared against infection count (*orange*). The solid orange line is the mean and shading the boundaries of the middle 95 percentile from 200 simulations and identical to **Figure 4A**.

## Algorithm 1: Sabin2 Intra-Host Molecular evolution

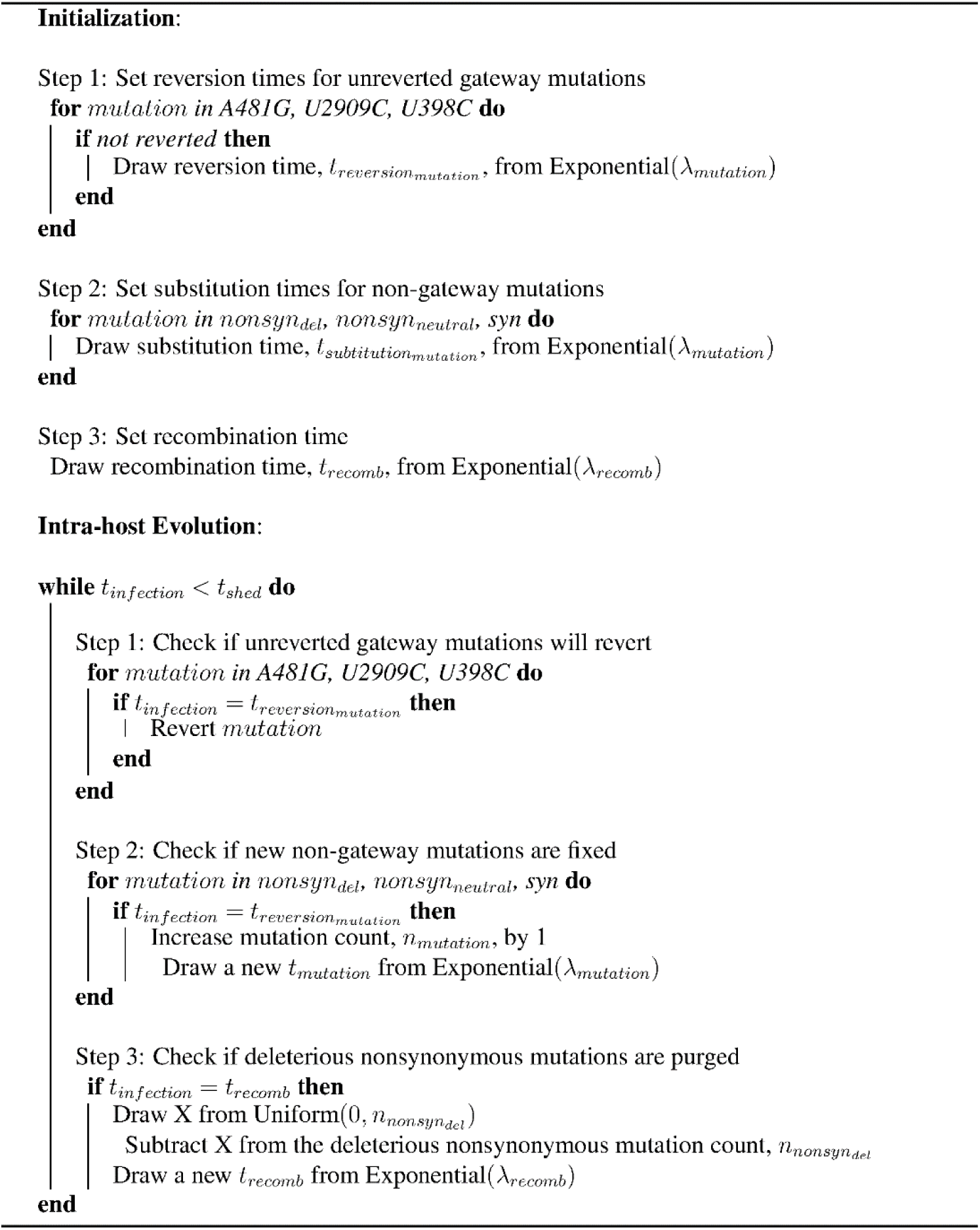

**Supplementary Figure 4.** Pseudocode for simulating Sabin 2 molecular evolution

**Supplementary Table 1.**
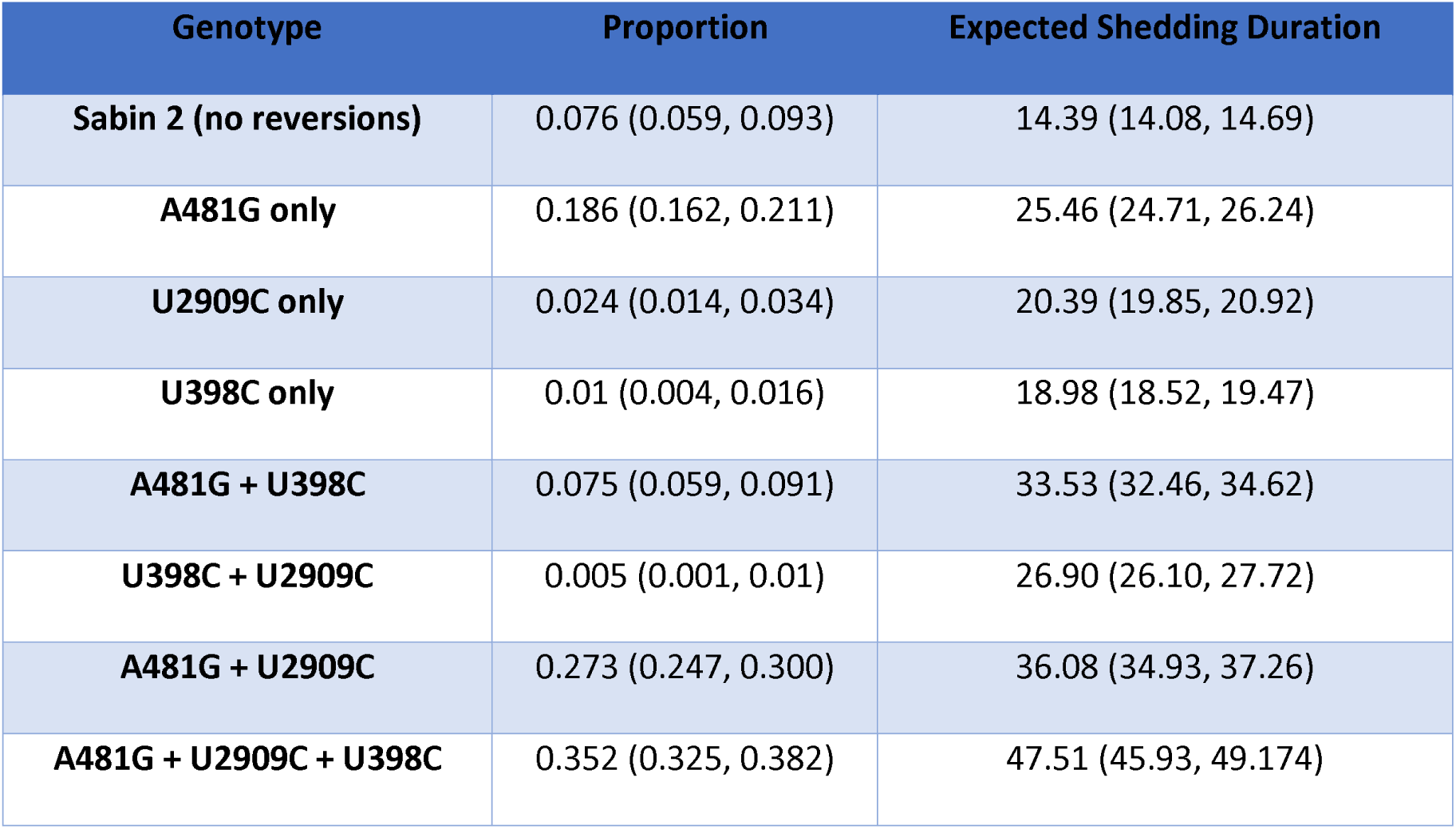
Final viral genotypes in primary vaccine recipients and the expected shedding durations of individuals infected with these genotypes, assuming no further reversion. Bootstrapped confidence interval for the mean were calculated from 1000 simulated primary vaccinations, repeated 1000 times. Shedding durations are reported in days. Sabin 2 in this table is defined as a viral genotype with zero gatekeeper reversions.

**Supplemental File 1** Accession numbers and study origins of the whole genome sequences

**Supplemental File 2** Accession numbers and study origins of the VP1 segments

## Notes

### Author Declarations

No approval necessary; only public and anonymous data were used

### Summary of Updates

Author order corrected; Manuscript title updated

