## Supplemental Information for "From Vaccine to Pathogen: Modeling Sabin 2 Vaccine Virus Reversion and Evolutionary Epidemiology"

Michael Famulare

Building IV, 3150 139th Ave SE, Bellevue, WA 98005

+1 (425) 691-3327

**SI Appendix**

*Calibrating Sabin 2 molecular evolution*

Simulated Sabin 2 molecular is based on the reversion of three gateway mutations (A481G, U2909C, U398C) as well as the accumulation of nonsynonymous mutations (deleterious and neutral) and synonymous mutations. Intra-host substitution rates (λ) were previously calculated from 241 VP1 segments from Type 2 Sabin-like poliovirus collected during routine acute flaccid paralysis surveillance in Nigeria from 2009-2012 (1, 2). These estimates were obtained by counting the frequency of the three gateway mutations and examining the accumulation of nonsynonymous and synonymous mutations.

We used the average, posterior intra-host substitution rates reported in the Nigeria study (1) for the three gateway mutations as their respective point estimates for intra-host substitution rates in our study ($\lambda_{A481G}$, $\lambda_{U2909C}$, $\lambda_{U398C}$) (**Appendix Table 1**). The synonymous substitution rate calculated from the Nigerian dataset (1) were not significantly different from those estimated from wild poliovirus (3). However, the nonsynonymous substitution rates inferred from the Nigerian dataset were two orders of magnitude higher than from WPV. This difference likely reflects a change in selection pressure as Sabin 2 due to changing epidemic circumstances and as the virus phenotypically approaches WPV. As a result, we could not directly use the nonsynonymous substitution rates reported from the Nigerian dataset.

*Nonsynonymous mutations: Neutral and Deleterious*

We divided the distribution of fitness effects into two categories, deleterious and neutral, and assign each with a separate intra-host substitution rate, $\lambda_{nonsyn, del}$ and $\lambda_{nonsyn, neutral}$. These rates were calibrated to the Sabin 2, Sabin 2-like, and cVDPV2 whole genome sequences and VP1 segments downloaded from Genbank (**Methods**). These sequences include the ones collected in the Nigerian study (1) but also include sequences reported to have been circulating for up to twenty years. Our goal was to infer $\lambda_{nonsyn, del}$, $\lambda_{nonsyn, neutral}$, $\lambda_{recombination}$, and $\lambda_{syn}$ based on the observed number of nonsynonymous mutations in the whole genome samples, the observed number of synonymous and nonsynonymous mutations in the VP1 segment samples (1).

We simulated Sabin 2 molecular evolution in the absence of transmission to calibrate these parameters. During each simulation, 1000 Sabin 2 genomes were evolved independently in daily time steps for twenty years. Mutation accumulation and reversion was modeled by drawing from an exponential distribution with the corresponding intra-host substitution rates. The average number of nonsynonymous mutations (deleterious and neutral) were recorded at each time step and compared to the data using a Poisson pseudo-log-likelihood function:

$$L= \sum_{i}^{n_{wgs}} (\hat{x}_{wgs,t_{i}}+\log\left( x_{i}! \right)-x_{i}log(\hat{x}_{wgs,t_{i}}))+ \sum_{i}^{n_{vp1}} (\frac{903}{7400}\hat{y}_{wgs,t_{i}}+\log\left( y_{i}! \right)-y_{j}log(\frac{903}{7400}\hat{y}_{wgs,t_{i}}))$$

(S1)

where $\hat{x}_{wgs,t_{i}}$ is the average number of nonsynonymous mutations in the simulated whole genome sequences at timepoint $t_{i}$, the time since vaccination for a given sample, $x_{i}$ is the number of nonsynonymous mutations observed for a given whole genome sequence,$\hat{y}_{wgs,t_{i}}$ is the average number of mutations (nonsynonymous and synonymous) mutations in the simulated VP1 segments at timepoint $t_{i}$, $y_{i}$ is the observed number of mutations a given VP1 segment, $n_{wgs}$ is the number of samples with whole genome sequences, and $n_{vp1}$ is the number of samples with vp1 sequence. Note that we assume that genome-wide mutation rates are the same in the VP1 segment, allowing us to calculate simulated VP1 mutations by multiplying $\hat{y}_{wgs,t_{i}}$ by the proportion of the genome within the VP1 segment. $t_{i}$ for each sample was calculated by dividing the observed number of synonymous mutations by $\lambda_{syn}$.

Posteriors for $\lambda_{nonsyn, del}$, $\lambda_{nonsyn, neutral}$, $\lambda_{recombination}$, and $\lambda_{syn}$ were generated using an Approximate Bayesian Computation - Sequential Monte Carlo (ABC-SMC) (4) with 300 particles and three iterations. Each particle tests a parameter set by independently running the simulation described above. Priors for$\lambda_{syn}$ , $\lambda_{nonsyn, del}$ and $\lambda_{nonsyn, neutral}$ were based on the Nigerian nonsynonymous substitution rates and the WPV nonsynonymous substitution rates, respectively (1). A prior for $\lambda_{recombination}$ was obtained by performing a parameter sweep using the priors for the other three parameters to identify a value that approximates the nonsynonymous mutations and VP1 mutations from samples collected within one year of vaccination (**Appendix Figure 1**). Error thresholds for each iteration were empirically generated by drawing 1000 parameter sets from our priors and set at the 30^th^, 10^th^, and 5^th^ percentiles. Priors and the final posterior estimates are reported in **Appendix Table 1**. The mean posterior value was used as our point estimates for all other simulations.

*Genotype to Phenotype: Calibrating shedding duration*

*Identifying selection coefficients and the base shedding duration*

The average shedding duration for poliovirus is lognormally distributed, with an average 30.3 days for Sabin 2 vaccine recipients and 43 days during WPV infection in immunologically naïve individuals(5). Here, we attribute the increase in shedding duration to the reversion of the three gateway mutations and a small number of deleterious nonsynonymous mutations. Our goal was to replicate the change in shedding duration from Sabin 2 and WPV as Sabin 2 evolves using **Equations 1-8**. We also utilized a modified form of **Equation 5**:

$$Duration={Duration}_{initial}*\left( e^{S_{dur, nonsyn} + Norm\left( 0,\sigma\right)\sqrt{s_{dur, nonsyn}}} \right)^{\Delta_{nonsyn,del}}$$

(S2)

where $\Delta_{nonsyn,del}$ is the total change in deleterious nonsynonymous mutations at each timestep due to either substitution or backmutation. The difference between Equation 5 and Equation S2 is the inclusion of a deleterious nonsynonymous mutation term that decreases shedding duration.

For the three gateway mutations, we assume that the selection coefficient is proportional to the relative rate of fixation compared to A481G, the fastest gateway mutation to fix (average 7 days). This allows us to redefine their selection coefficients using a single selection coefficient ($s_{dur, point}$):

$$s_{inf, gateway}=s_{dur,point}*\frac{\log\left( \lambda_{gateway} \right)}{\log\left( \lambda_{A481G} \right)}$$

(S3)

Note that $s_{dur,point}$ is equivalent to $s_{dur,A481G}$. Relating $s_{gateway}$ to the relative ratio of log fixation rates was decided empirically and resulted in better calibration fits. We calibrated simulated shedding durations using four parameters: $s_{dur,point}$, $s_{dur,nonsyn_{del}}$, $\mu_{S2}$ (the base shedding duration of unevolved Sabin 2, $G_{0,0,0,0,0,0}$) and $\sigma$ the standard deviation in shedding duration. To calibrate these three parameters, we utilized two sets of simulations: one where we replicate the shedding duration of Sabin 2 vaccinees and one where we replicate the shedding duration of WPV infected individuals. Each simulation simulates 1000 infections and allows the shedding duration to be affected by gateway mutation reversion and deleterious nonsynonymous mutation accumulation.

We simulated 1000 Sabin 2 vaccinations in immunologically naïve individuals (Nab = 1) and allowed newly acquired mutations to extend the shedding duration as defined by **Equation S2**. Once all vaccinated individuals stop shedding, we compared simulated shedding durations against 1000 samples from the lognormal distribution describing the shedding durations following clinical Sabin 2 vaccination (mean = 30.3, std = 1.86) (5). We also simulated WPV shedding durations by infecting 1000 immunologically naïve individuals with $G_{1,1,1,9. X,X}$. This genome represents the average, phenotypically relevant genotype of fully reverted cVDPV2 sequences. Note that we assume nine deleterious nonsynonymous mutations because it represents the average steady-state value in our simulated genomes after all intra-host substitution rates were calibrated. The number of neutral nonsynonymous mutations and synonymous are denoted by X and assigned an arbitrary value during simulations because they do not affect shedding duration. These shedding durations were compared against 1000 samples from the lognormal distribution describing shedding duration in natural WPV infections (mean = 43.0, std = 1.69) (5).

Posteriors for $\mu$, $s_{dur,point}$,and $s_{dur,nonsyn_{del}}$ were generated using ABC-SMC with 300 particles and three iterations. Priors were obtained by running a simplified version of the simulations described above, where we 1) assume mutations always arise at their average fixation times, 2) assume that WPV does not acquire additional nonsynonymous mutations (ie $G_{1,1,1,9. X,X}$remains constant), 3) obtain parameter estimates by minimizing the least-squares difference between simulated and observed means. For the ABC-SMC, distance was defined using a pseudo-log likelihood function:

$$L= -\frac{n}{2}\ln\left( 2\pi\right)-\frac{n}{2}\ln\left( \hat{v} \right)-\frac{1}{2\sigma^{2}}\sum_{j-1}^{n} \left( x_{j}-\hat{m} \right)^{2}$$

(S4)

where $x_{j}$ denotes a random sampling form the target lognormal distribution, n is the total number of samples made, $\hat{m}$ is the mean and $\hat{v}$ is the variance of the simulated shedding durations. Error thresholds for each iteration were empirically generated by drawing 1000 parameter sets from our priors and set at the 30^th^, 10^th^, and 5^th^ percentiles.

Our point estimate for $s_{nonsyn_{del}}$ was -0.006 (-0.04, 0.03) but is too small to have a noticeable effect on shedding duration (**Appendix Figure 2**). Overall, there was insufficient evidence to definitively identify $s_{nonsyn_{del}}$ as increasing or decreasing shedding duration, as the posterior occupies both positive and negative values. As a result, we simplified our final shedding duration model and assert $s_{nonsyn_{del}}$ as zero (equivalent to **Equation 5**). Final calibration estimates are in **Appendix Table 2**.

*Identifying the impact of immunity on shedding duration*

Thus far, our calibration has focused on the shedding duration in immunologically naïve individuals. To calibrate shedding duration in individuals, we also needed to re-evaluate $\delta$, the parameter that governs the decrease in shedding duration due to immunity (**Equation X**). To calibrate $\delta$, we simulated the expected shedding durations for individuals with $N_{ab}$ ranging from 1-4096. Using all previously calibrated parameter estimates, we used ABC-SMC with 300 particles and three iterations to compare simulated shedding durations with the expected shedding durations for Sabin 2 reported in (5) for each $N_{ab}$ titer examined. A prior was obtained by performing a parameter sweep across $\delta$, ranging from 0-2; our previous estimate for $\delta$ was 1.16 (5). Distance was calculated as the least square difference in average shedding duration. Error thresholds for each iteration were empirically generated by drawing 1000 parameter sets from our priors and set at the 30^th^, 10^th^, and 5^th^ percentiles. Priors and the final posterior estimates are reported in **Appendix Table 3**.

*Genotype to Phenotype: Calibrating Infectiousness*

WPV is approximately four times more infectious than Sabin 2 (2.3 CCID50 units for WPV and 8 CCID50 units for Sabin 2). As with shedding duration, we attribute this increase in infectiousness (decrease in CCID50 units) to the acquisition of the three gateway mutations and the accumulation of deleterious nonsynonymous mutations. This is governed by $\beta_{g}$ (**Equation 8**), which describes the change in infectiousness associated with each mutation. Our goal was to infer $s_{inf,A481G}$, $s_{inf,U2909C}$, $s_{inf,U398C}$, and $s_{inf, nonsyn_{del}}$.

Like with shedding duration, we simplify parameter calibration by assuming the selection coefficients for each of the gateway mutations is proportional to their relative fixation rates:

$$s_{inf, gateway}=s_{inf,point}*\frac{\lambda_{gateway}}{\lambda_{A481G}}$$

(S5)

Note that the fixation rate ratios are not in log space. Infectiousness is determined only at the onset of infection. Point estimates were obtained by minimizing **Equation S6** using a numerical approximation method (scipy.optimize.minimize, method = ‘TNC’):

$$\beta_{wpv}=\frac{\beta}{\left( 1+s_{inf,A481G} \right)^{1} \left( 1+s_{inf,U2909C} \right)^{1} \left( 1+s_{inf,U398C} \right)^{1} \left( 1+s_{inf,nonsyn_{del}} \right)^{9}}$$

(S6)

where $\beta$ and $\beta_{wpv}$ are substituted with the average estimates obtained from various OPV trials (5). Note that the WPV genotype is again defined as $G_{1,1,1,9. X,X}$. Point estimates are provided in **Appendix Table 4**.

*Calibration to mOPV2 clinical trial data*

We previously developed a multiscale, household demographic transmission model (**cite Matlab paper**) that simulates changing poliovirus immunity and transmission with different contact rates across different demographic scales: 1) household, 2) bari, an intergenerational living arrangement unique to rural Bangladesh, 3) village, and 4) region. Contact rates and fecal-oral doses were calibrated to Sabin 2 shedding following a monovalent, Sabin 2 Oral Poliovirus (mOPV2) campaign performed in Matlab, Bangladesh. After incorporating Sabin 2 evolution, we recalibrated our multiscale transmission model by performing a parameter sweep across the fecal-oral dose parameter. All other parameters regarding demographics and contact rates were not recalibrated. Our estimate for the fecal oral dose was 3.6e-7 (2.6e-7, 5.1e-7), which is about an order of magnitude lower than our previous estimate (2.5e-6) (**cite Matlab paper**).

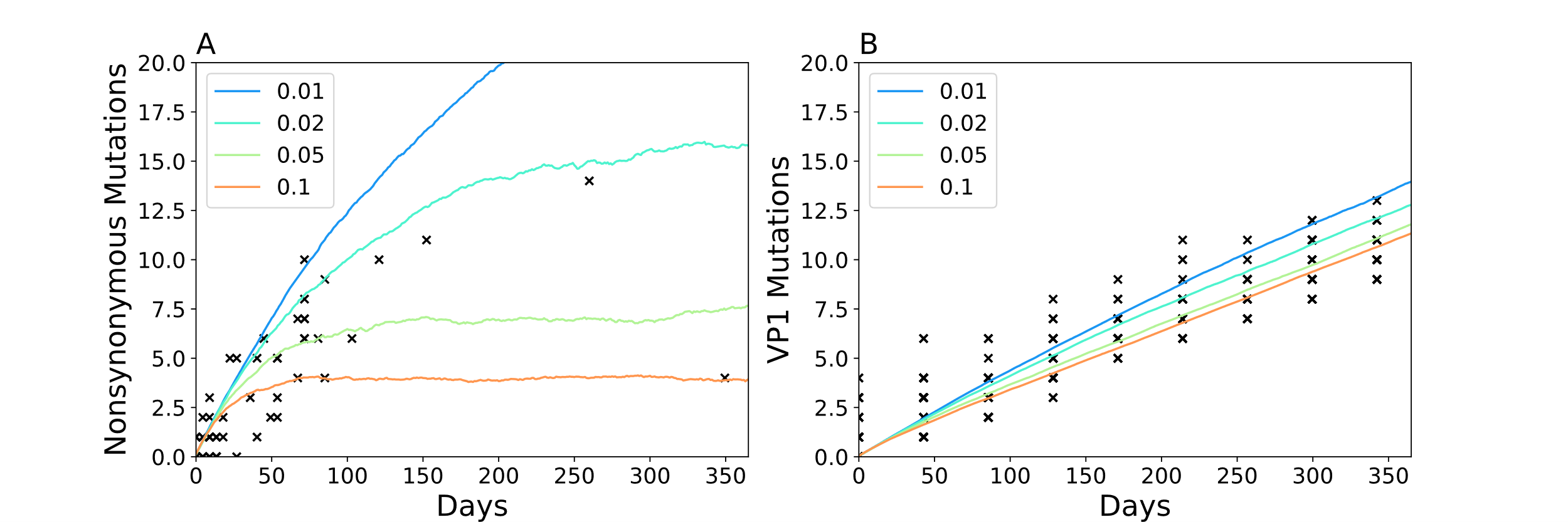

**Appendix Figure 1** $\lambda_{recomb}$parameter sweep using the $\lambda_{nonsyn, del}$, $\lambda_{nonsyn, neutral}$, and $\lambda_{syn}$ priors obtained form the Nigeria study compared against **A)** nonsynonymous mutations in the whole genome sequences and **B**) VP1 segments. $\lambda_{recomb}$ is measured as the average number of recombination events per day and the time between recombination events is assumed to be exponentially distributed. Each line is the mean number of mutations from 1000 evolving genomes. Black x’s are the data.

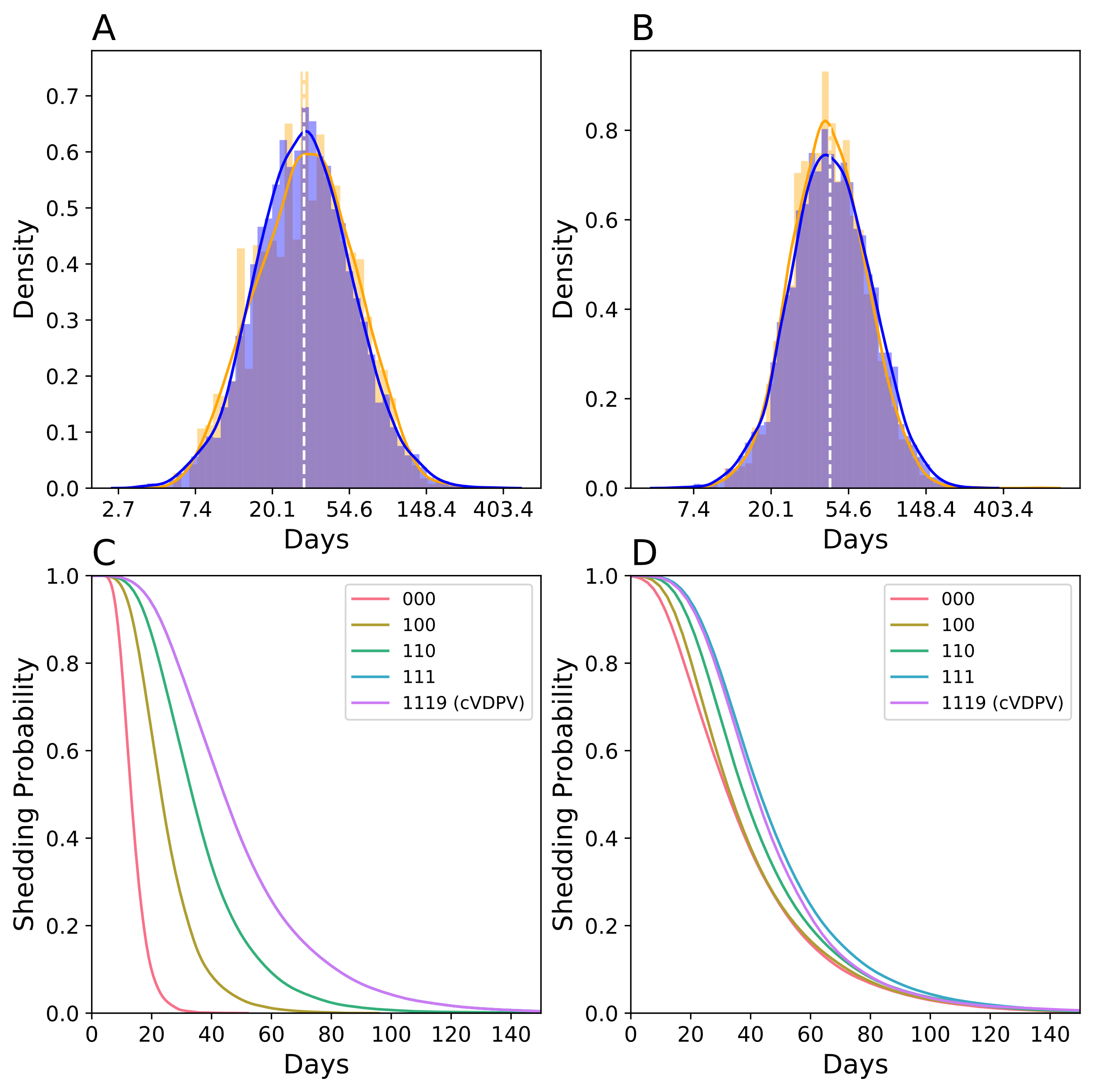

**Appendix Figure 2** Shedding duration observed in **A)** Sabin 2 primary vaccination recipients **B)** WPV infections. For **A** and **B**, simulated shedding durations (*orange*) are compared against the lognormal distribution describing clinically observed shedding durations (*blue*). Dotted white line denotes the expected mean shedding duration times following Sabin 2 vaccination and WPV infections. Shedding durations of different strains (red = $G_{0,0,0,0,X,X}$, yellow = $G_{1,0,0,0,X,X}$, green = $G_{1,1,0,0,X,X}$, blue = $G_{1,1,1,0,X,X}$, pink = $G_{1,1,1,9,X,X}$) assuming static genotypes with no evolution (**C**) and assuming mutations continue to be acquired throughout the infection (**D**). Simulations assume deleterious nonsynonymous mutations effect on shedding duration ($s_{dur,nonsyn_{del}}$ = -0.006).

| **Parameter** | **Prior** | **Mean Posterior** | **Lower 95%** | **Upper 95%** |
| --- | --- | --- | --- | --- |
| λ_recomb_ | 3.62E-06 | 5.24E-06 | 3.13E-06 | 7.50E-06 |
| λ_nonsyn,del_ | 2.05E-05 | 1.83E-05 | 9.78E-06 | 3.02E-05 |
| λ_nonsyn,neutral_ | 3.32E-06 | 3.34E-06 | 2.93E-06 | 3.66E-06 |
| λ_syn_ | 3.00E-05 | 3.16E-05 | 3.02E-05 | 3.29E-05 |
| λ_A481G_ | 1.54E-01 | 1.54E-01 | 7.00E-02 | 2.97E-01 |
| λ_U2909C_ | 4.70E-02 | 4.70E-02 | 1.80E-02 | 9.60E-02 |
| λ_U398C_ | 2.10E-02 | 2.10E-02 | 6.00E-03 | 4.80E-02 |

**Appendix Table 1 Intra-host substitution rates** (substitutions per base pair per day) All priors are based on VP1 segments collected in Nigeria. The mean posterior value is used as our point estimate. Posterior estimates for the three gateway mutations are identical to the values reported in the Nigeria study (1).

| **Parameter** | **Prior** | **Posterior** | **Lower 95%** | **Upper 95%** |
| --- | --- | --- | --- | --- |
| μ | 12.000 | 13.160 | 11.001 | 15.394 |
| σ | 0.300 | 0.325 | 0.274 | 0.378 |
| s_dur,A481G_ | 0.600 | 0.574 | 0.389 | 0.757 |
| s_dur,U2909C_ | 0.367 | 0.351 | 0.238 | 0.463 |
| s_dur,U398C_ | 0.291 | 0.278 | 0.188 | 0.367 |
| *s_dur,nonsyn,del_* | *0.000* | *-0.006* | *-0.041* | *0.029* |
| δ |  |  |  |  |

**Appendix Table 2 Shedding duration parameters** assuming $s_{dur,nonsyn_{del}}$affected shedding duration. *δ* was not estimated for this version of the model.

| **Parameter** | **Prior** | **Posterior** | **Lower 95%** | **Upper 95%** |
| --- | --- | --- | --- | --- |
| μ | 12.000 | 13.790 | 11.990 | 14.410 |
| σ | 0.300 | 0.360 | 0.310 | 0.400 |
| s_dur,A481G_ | 0.600 | 0.535 | 0.460 | 0.660 |
| s_dur,U2909C_ | 0.367 | 0.327 | 0.281 | 0.404 |
| s_dur,U398C_ | 0.291 | 0.259 | 0.223 | 0.320 |
| s_dur,nonsyn,del_ | 0.000 | -0.050 | -0.080 | -0.012 |

**Appendix Table 3 Shedding duration parameters** assuming $s_{dur,nonsyn_{del}}$had no effect on shedding duration phenotype. These were the final parameter estimates used in our model.

| **Parameter** | **Point Estimate** |
| --- | --- |
| β_wpv_ | 2.30 |
| β | 8.00 |
| s_inf,A481G_ | 1.82 |
| s_inf,U2909C_ | 0.56 |
| s_inf,U398C_ | 0.25 |
| s_inf,nonsyn,del_ | -0.05 |

**Appendix Table 4 Infectiousness parameters**. β_wpv_ and β_S2_ were obtained from (5).
